# Healthcare professionals’ collaboration and satisfaction within an innovative primary care network for patients with chronic musculoskeletal pain: a mixed method study

**DOI:** 10.64898/2026.03.23.26349104

**Authors:** Cynthia Lamper, Mariëlle Kroese, Marion de Mooij, Jeanine Verbunt, Ivan Huijnen

**Affiliations:** Department of Rehabilitation Medicine, Functioning, Participation & Rehabilitation, Care and Public Health Research Institute (CAPHRI), Faculty of Health, Medicine and Life Sciences, Maastricht University, Maastricht, the Netherlands; Department of Health Services Research, Care and Public Health Research Institute (CAPHRI), Faculty of Health, Medicine and Life Sciences, Maastricht University, Maastricht, the Netherlands; Centre of Expertise in Rehabilitation and Audiology, Adelante, Hoensbroek, the Netherlands

**Keywords:** Interprofessional collaboration practice, joy and meaning in work, Quadruple Aim, primary care, chronic musculoskeletal pain

## Abstract

**Background/Objective:** The Network Pain Rehabilitation Limburg (NPRL) was established to provide integrated, biopsychosocial-based rehabilitation care for patients with chronic musculoskeletal pain, emphasizing the delivery of appropriate care by the right person at the right place and cost. This study examines the perceived interprofessional collaboration practice (ICP) and work satisfaction among primary care healthcare professionals engaged in NPRL.

**Patients and Methods:** A mixed-methods approach involved seven general practitioners (GPs), twenty-four therapists (physiotherapists and occupational therapists), and five mental health practice nurses in eleven semi-structured focus groups and one interview conducted from 2017 to 2020. The Interprofessional Collaboration Attainment Survey quantitatively measured healthcare professionals’ ICP abilities before and after NPRL participation. Qualitative analysis, structured around existing ICP frameworks and the Quadruple Aim, was based on interview data.

**Results:** Findings revealed stable ICP and work satisfaction, with discussions focusing on transitioning to a biopsychosocial perspective on chronic pain and its implications, along with concerns about GP burden and insurer reimbursement issues. Significant enhancements were noted in communication and team functioning (p < 0.05).

**Conclusions:** Overall, healthcare professionals reported positive experiences with NPRL’s integrated approach, showcasing dedication to providing rehabilitation care for chronic musculoskeletal pain in primary care. Recommendations for improving ICP included advocating for a broader societal biopsychosocial view of chronic pain, introducing case managers in primary care to support GPs, and exploring alternative reimbursement models with insurers. However, significant transformations to impact work satisfaction and ICP may necessitate more time and consideration.

**Key messages:**

- Healthcare professionals were positive and demonstrated commitment to interdisciplinary collaborations in primary care for guidance of patients with chronic musculoskeletal pain.
- Facilitators of interprofessional collaboration and increased work satisfaction could include a more biopsychosocial view of chronic musculoskeletal pain in society, reducing the burden on GPs by introducing a case manager in primary care, and implementing a different reimbursement model for healthcare professionals by insurers.
- Healthcare professionals expect that the benefits in terms of interprofessional collaboration practice and work satisfaction will become increasingly visible on the long term.

## Introduction

In Europe, 19% of the population suffers from chronic musculoskeletal pain (CMP) [1]. With an aging population, this number is expected to increase. CMP, with varying duration and intensity of complaints, considerably impacts wellbeing. For example, low back pain accounts for many years lived with disability, even more than conditions such as COPD, diabetes, and major depression [2,3].

Accordingly to the World Health Organization (WHO), rehabilitation is an essential component of integrated health services [4]. Rehabilitation can be offered as either a monodisciplinary or multidisciplinary treatment in primary care or within specialized care. By mapping the influencing and maintaining factors from an integral perspective, the right care can be delivered in the right place [5,6]. This means that patients with low to moderate complexity of chronic musculoskeletal pain may benefit from monodisciplinary treatment, such as advice or reactivation. In contrast, patients with more complex CMP complaints may benefit from a biopsychosocial approach delivered by an interdisciplinary team of healthcare professionals (HCPs) from different settings and disciplines.

However, the transition towards integrated health services increases pressure on HCPs, requiring more time and commitment without extra pay. Implementation changes in healthcare is challenging, especially since current work pressure and burnout rates are already high in Dutch healthcare [7–9]. These factors significantly impede professionalism and quality of care, while increase the risk of staff turnover and medical errors [10–14]. Given these challenges and the need for workforce engagement in healthcare transitions, work satisfaction is of high interest [14,15].

To address these issues, one way to tackle the complex health needs of populations and patients is by improving Interprofessional Collaborative Practice (ICP) [16,17]. The increasingly complex health needs of the population and patients can only be met by promoting ICP. The WHO (2010) defines collaborative practice as occurring *“when multiple health workers from different professional backgrounds provide comprehensive services by working with patients, their families, carers, and communities to deliver the highest quality of care across settings”* [18]. Interprofessional Collaborative Practice in rehabilitation is not commonly applied but is essential for providing comprehensive and effective care [19]. Key factors that support IPC include strong team culture, mutual trust, clear communication, and well-defined roles [20–23]. However, barriers such as organizational constraints and lack of clarity in IPC terms need to be addressed [24,25]. Rehabilitation care is transitioning towards working in integrated health services, such as integrated in primary care, in which healthcare professionals work following ICP [5,17].

Currently, the Dutch healthcare for patients with CMP is not organized as an integrated health service. The organization is fragmented and a common vision regarding the content and place of treatment is lacking. Therefore, the Standard of Care for patients with CMP was introduced to enhance healthcare transition aiming to deliver integrated, biopsychosocial based rehabilitation care with the right care, at the right place, by the right person, for the right price to improve patients’ level of functioning [26,27]. In 2017, based on this standard, the Network Pain Rehabilitation Limburg (NPRL) was developed [28]. The aim was to provide integrated care with a biopsychosocial approach for patients with CMP in order to improve their level of functioning. Healthcare ranges from advice-only and monodisciplinary treatments to interdisciplinary treatments depending on the complexity of patients’ pain complaints. Interprofessional teams consist of general practitioners (GPs), practice nurses mental health [Dutch job title], and therapists (physiotherapists and occupational therapists) in primary care. They receive interdisciplinary education in order to reach common vision, language, communication and well-defined roles. Collaboration will be supported by facilitating communication between patients and all HCPs involved in the trajectory of an individual patient by E-health. Additionally, teams held scheduled meetings at least every six weeks to discuss patients, or sooner when needed. This ensured a common understanding of the biopsychosocial approach and rehabilitation treatment. A first version of the network (NPRL1.0) was evaluated in a feasibility study (2017-2018). These findings led to refinements resulting in NPRL2.0, which was evaluated by a (cost)-effectiveness study (2019-2020). More background information on the development and content of NPRL can be found in previously published studies [26–28].

Since this approach to care organization is new in the Netherlands, it is important to understand how implementing a new integrated care model affects perceived work satisfaction and ICP of HCPs. This study aims to describe perceived ICP and work satisfaction of primary care HCPs participating in NPRL.

## Methods

### Design and data collection

This mixed-methods study used both qualitative and quantitative data on ICP and work satisfaction, namely, the transcripts of focus groups conducted during the NPRL 1.0 study, and focus groups and a quantitative survey related to NPRL2.0. See figure 1 for schematic overview of the data collection.

**Figure 1:**
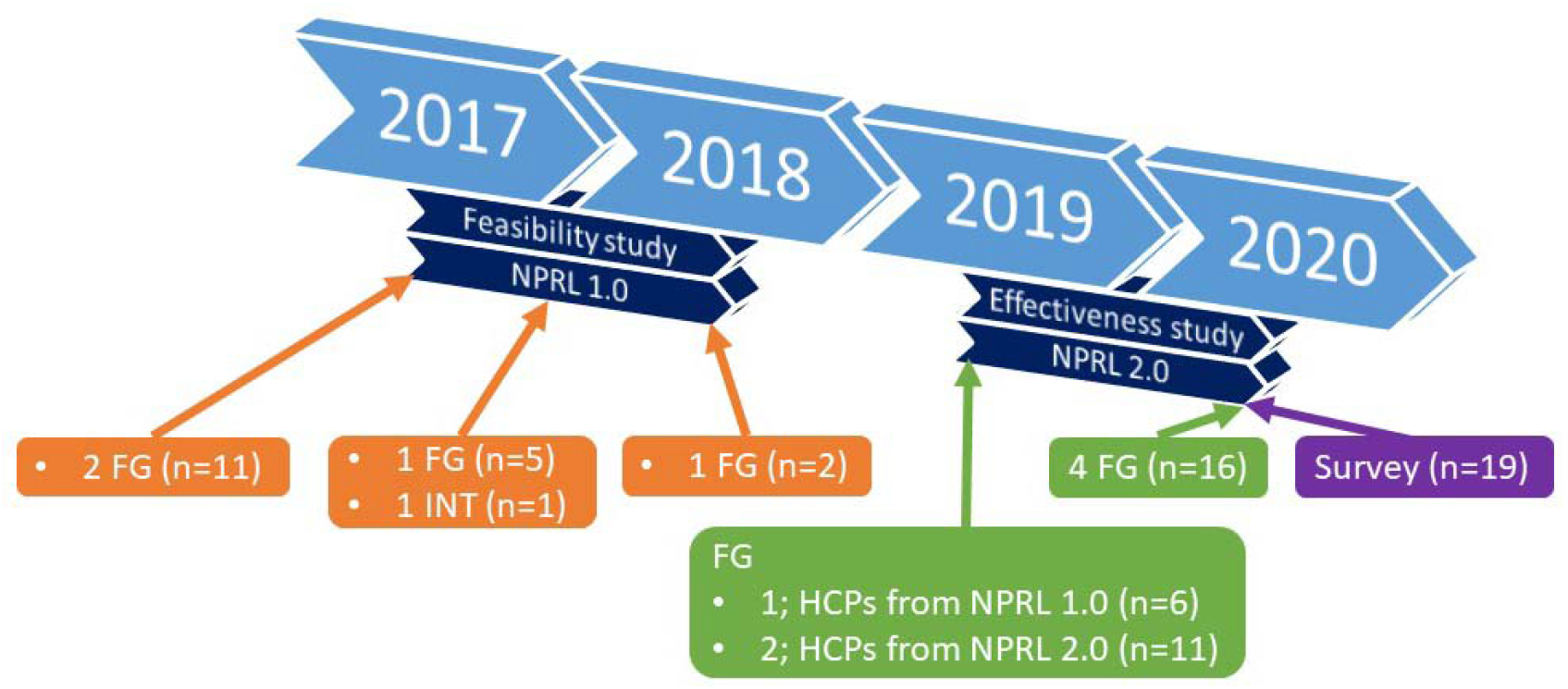
Overview of data collection of feasibility study in NPRL 1.0, and effectiveness study in NPRL2.0. FG: focus group, INT: interview, HCP: healthcare professional.

### Participants

Sixty-six HCPs who provided IPC in NPRL were asked to participate in the focus groups and survey. Among the participants were 23 general practitioners, 31 therapists (physiotherapists and occupational therapists), and 12 practice nurses mental health. The inclusion criterion was participating the educational meetings of NPRL. Informed consent for participation in this study was obtained.

### Theoretical frameworks

Interviews and qualitative analysis were structured based on existing frameworks: 1) Conceptual Framework for ICP, 2) Job-Demand-Control-Support Model, and 3) Quadruple Aim (figure 1, Explanation of the constructs: Appendix A) (14, 23, 24). They are combined to ‘the Integrated ICP and Quadruple Aim framework’.

#### Conceptual Framework for Interprofessional Collaborative Practice (ICP)

Stutsky and Spence Laschinger [2014] conceptualized ICP as a four-dimensional construct including understanding of roles, interdependence, knowledge exchange, and collective ownership of goals [29]. The ICP framework can be used as a guide for facilitating ICP in organizations and networks to enhance patients’ safety and quality. It can also facilitate the awareness of HCPs of individual attitudes and behaviors and team interactions to improve patient safety and quality.

#### Job-Demand-Control-Support Model

Joy and meaning in work are influenced by the occurrence of mental strain in the work environment. The Job-Demand-Control-Support Model explains how job characteristics influence employees’ psychological well-being [30,31]. The model describes that work situations with high demands lead to work stress. The stress can be decreased by gaining great job control and developing strong social support from colleagues and supervisors. If a HCP has both low control and low social support it can eventually lead to harmful health outcomes

#### Quadruple Aim

The Quadruple Aim is an approach to optimize health system performance, proposing that healthcare institutions simultaneously pursue four aims. The first three aims (improving population health, reducing costs, and enhancing patient experience of care) provide a rationale for the existence of a health system. The fourth aim, improving the work-life of health care providers, becomes a foundational element for the other goals to be realized [14,15]. The focus of this study is on work satisfaction. Sikka et al. [2015] define it as creating the conditions for the healthcare workforce to find joy and meaning in their work and in doing so, improving the experience of providing care [15]. According to them, meaning in work is a sense of importance in daily work. Joy in work is the feeling of success and fulfillment that results from meaningful work. Joy and meaning in work are positively related to work satisfaction [32]. Work behaviors and attitudes of ICP are part of this fourth Aim. In our Integrated ICP and Quadruple Aim framework, behavior, attitudes, and work satisfaction are connected.

### Data collection

The current mixed methods study used both qualitative and quantitative data on ICP and work satisfaction collected in two studies in the context of NPRL. All HCPs involved in NPRL1.0 and NPRL2.0 were asked to participate in this study, ensuring that each individual HCP participated at least once in a focus group or interview.

#### Focus groups and interview

During the feasibility study of (NPRL1.0), focus group 1 and 2 were performed in person in December 2017; focus group 3 and the interview with a practice nurses mental health (who was not able to participate in the focus group) in May 2018; focus group 4 in October 2018. Although the subject of these focus groups and the interview was not specifically ICP and work satisfaction, this topic arose during the discussions (Table 1, Supplementary file 2). A document analysis was performed on transcripts of these four focus groups and one interview [33].

**Table 1:**
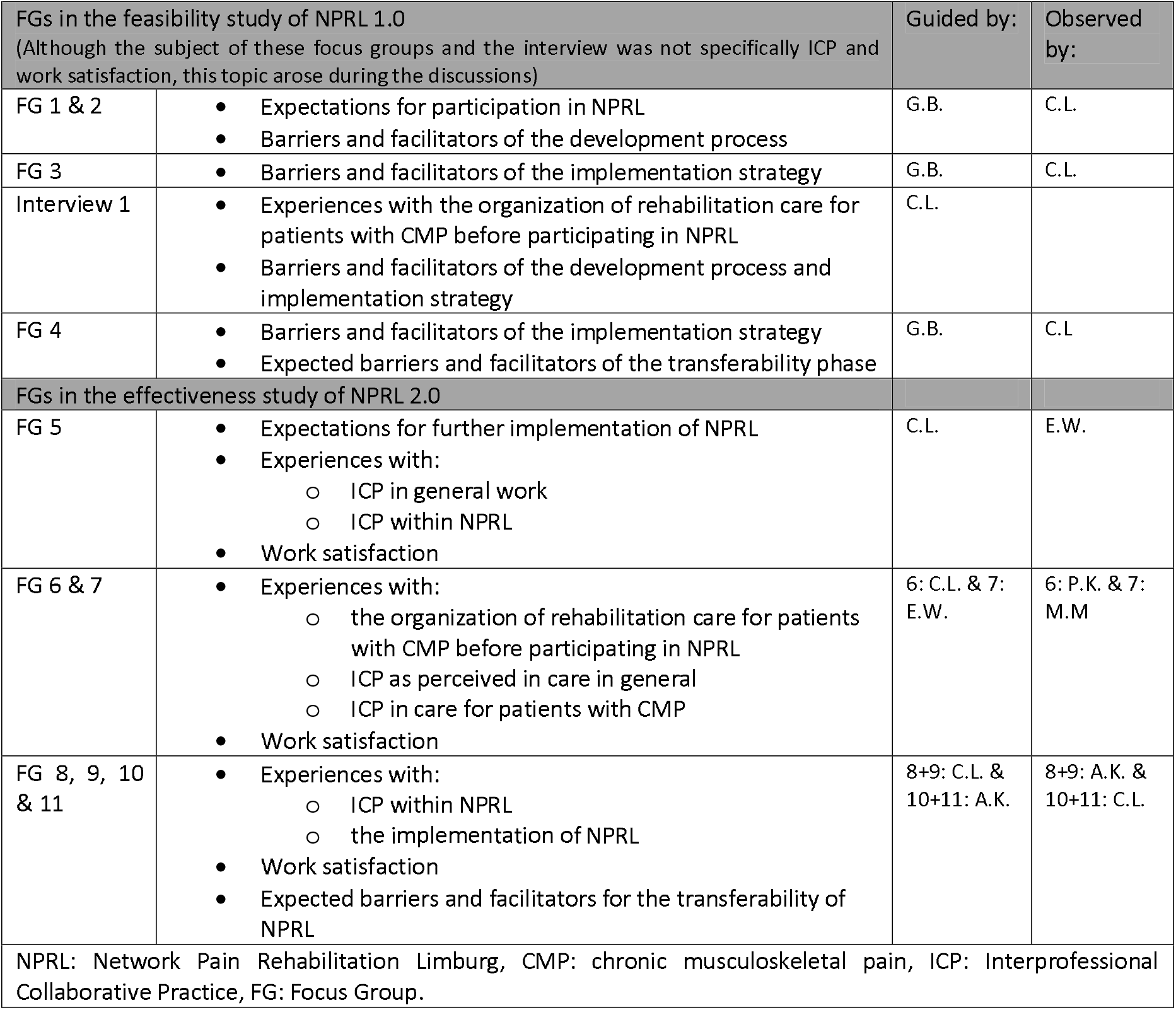
Content of FGs and interview.

In April 2019, three focus groups were conducted in person for the effectiveness study (NPRL2.0). Focus group 5 comprised HCPs who also participated in the feasibility study (NPRL1.0) and additionally, experiences of the previous 1.5 years were evaluated. Focus group 6 and 7 comprised only recently involved/enrolled HCPs of NPRL2.0. During the focus groups performed ICP and work satisfaction were specific topics (Table 1).

In April 2020, focus groups 8, 9, 10 and 11 were held with all involved HCPs for the effectiveness study (NPRL2.0). Due to COVID-19, these were performed with Zoom Video Communications (V5.0.2). During the focus groups performed ICP, work satisfaction, and expected barriers were specific topics (Table 1).

#### Survey on interprofessional collaboration practice competencies

The Interprofessional Collaboration Competencies Attainment Survey (ICCAS) was used to retrospectively measure HCPs’ ability to perform interprofessional competency skills before and after participating in NPRL [34–37]. The ICCAS is a 20-item survey with a 5-point scale from poor (1) to excellent (5), measuring six constructs: communication, collaboration, roles and responsibilities, collaborative patient-family centered approach, conflict management/ resolution, and team functioning. The ICCAS is validated in English and translated into Dutch for this study [35,36]. The survey was sent to all HCPs working in NPRL (n=37) by Qualtrics Survey Software (Qualtrics, Provo, UT) in April 2020.

### Data Analysis

Transcripts of the focus groups and surveys open questions were analyzed with NVivo software (NVivo V12 Pro). Codes were developed deductively from ‘the Integrated ICP and Quadruple Aim framework’ (figure 1) and inductively from the data. Two researchers (C.L. and M.M.) coded all the transcripts to ensure no themes were missed in the analysis. When disagreements about concepts arose, these were discussed until an agreement was reached.

Several themes were applied to various domains of the proposed Integrated ICP and Quadruple Aim framework. To avoid repetition of themes in multiple domains, each theme is solely described under the most important domain. In Figure 2, the presented domains are shown in bold and they are also presented in table 2.

**Table 2:** presented domains and themes.

| Table 2: presented domains and themes |  |
| --- | --- |
| Personal factors | Beliefs in Interprofessional Collaboration |
|  | • Trust<br>• Communication skills |
| Situational factors | • Leadership<br>• Support structure |
| Interprofessional Collaborative Practice | • Collective ownership of goals<br>• Knowledge exchange<br>• Understanding of roles |
| Work behaviors & attitudes | • Work satisfaction (joy and meaning)<br>• Work stress |
| Patient outcomes | • Biopsychosocial outcomes<br>• Satisfaction |
| Organizational outcomes | • Length of treatment |

**Figure 2.**
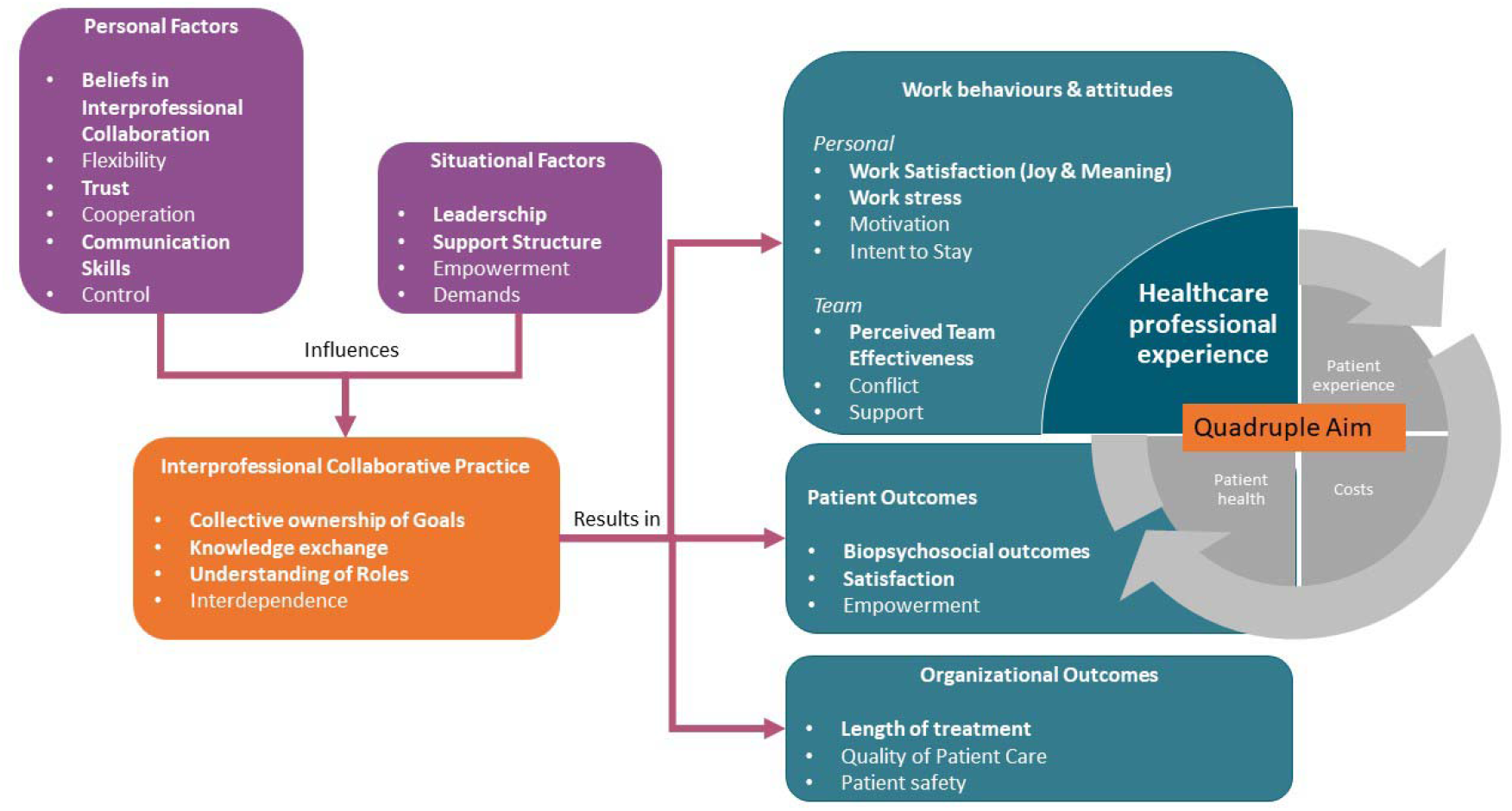
The Integrated ICP and Quadruple Aim framework used in this study is based on the Conceptual Framework for Interprofessional Collaborative Practice (ICP), Job-Demand-Control-Support Model, and Quadruple Aim. An explanation of the constructs can be found in Appendix A.

Data of the ICCAS were analyzed using descriptive statistics for quantitative variables and the signtest for changes in pre-test to post-test scores. The magnitude of this change was analyzed using Cohen’s d in IBM SPSS Statistics (V.24) [38]. Large differences were interpreted from scores > 0.8, medium differences from 0.80-0.50, small differences from 0.5-0.20, and negligible from <0.20 (37).

### Ethical Considerations

Ethical approval was obtained from the Medical Ethics Committee Z, the Netherlands, ethical approval numbers: METC17N133 and METCZ20190037. This study adheres to the Declaration of Helsinki. Written informed consent for participation in this study was obtained from HCPs.

## Results

In this study, a total of 36 HCPs participated, including 7 GPs, 24 therapists, and 5 mental health practice nurses (see Supplementary File 1). Twelve of these HCPs had also participated in NPRL1.0 (Focus Groups 1–4 and Interview 1), comprising 3 GPs, 8 therapists, and 1 mental health practice nurse. In NPRL2.0 (Focus Groups 5–10), 27 HCPs took part, including 5 GPs, 19 therapists, and 3 mental health practice nurses. Nineteen participants completed the first questionnaire (Q1), including 16 men and 21 women. Their years of work experience ranged from 0.5 to 34 years.

### Document analysis and focus groups

#### Personal factors

##### Beliefs in interprofessional collaboration practice

Many HCPs support ICP. Some noted existing effective ICPs for other diseases in their primary healthcare centers. For patients with CMP, optimizing ICPs and increasing interdisciplinary collaboration were desired. Reasons for HCPs to participate in NPRL included a desire to enhance ICPs, affinity with the patient population, and commitment to deliver efficient care. P31 expressed this sentiment: *“The reason I participate is to improve care quality*.*”* Some HCPs focused on CMP, others were drawn to the patient population, or sought to learn about motivating patients for biopsychosocial treatment. Before NPRL1.0, they viewed patients with CMP as complex due to subjective pain and common comorbidities. After participation, they still perceived these patients complex, but felt better equipped with tools and treatment options, thanks to training, giving them a greater sense of control. Some HCPs saw participation as an opportunity to showcase their innovative nature to patients.

##### Trust

Before NPRL1.0, HCPs lacked insights into each other’s disciplines and treatments. During the study, interprofessional educational sessions improved trust in each other’s competencies and treatment approaches. GPs felt confident when referring patients to participating therapists. During the study, not all HCPs were proactive in their participation. To optimize future collaboration, a more proactive attitude among some HCPs is recommended. Participating HCPs emphasized that new participants must prioritize care transformation for patients with CMP to effectively engage in NPRL2.0.

##### Communication skills

Before NPRL1.0, HCPs struggled to get interdisciplinary connections and sought more empowerment and support to facilitate interdisciplinary communication. As P34 noted: *“For patients, it is important to know where I refer someone to and the vision regarding CMP of that HCP or practice. And for me, it is good to note that a common vision is an aim of NPRL1*.*0*.*”* Bi-directional communication between GPs, therapists, and secondary or tertiary care was lacking when patients were referred, often resulting in incomplete treatment reports. During NPRL 1.0 and NPRL 2.0, most HCPs reported that the ease and effectiveness of communication had been maintained or improved. GPs had provided pain education more frequently and had explained biopsychosocial treatment to patients before referral to therapists. Some therapists felt supported by their GPs within the local network as the GPs prepared the patients for a biopsychosocial treatment before therapists’ consultations. HCPs anticipate that these streamlined communication channels will enhance collaboration in the future, with preference given to face-to-face contact over telephone communication.

#### Situational factors

##### Leadership

At NPRL1.0’s outset, HCPs viewed the NPRL project team as leaders, confident in successful execution due to dedicated staff from a regional rehabilitation center specialized in interdisciplinary pain rehabilitation treatment, managing the project. They believed in its potential because of clear boundaries and treatment guidelines. HCPs hoped the current project team would continue leading NPRL. P28 highlighted that reminders, organization guidance, and patient case education by the project team helped to maintain focus. Leadership varied across local networks, with GPs often seen as best suited to lead ICP due to their patient contact role. The shift from HCP as leader instead of the project management group is mentioned as important for continuing interdisciplinary consultations.

##### Support structures: financial support

Firstly, HCPs reported that their capacity and willingness to begin treatment for patients with inadequate health insurance coverage was limited.

Secondly, primary care HCPs indicate that a lack of financial compensation for interdisciplinary consultations and project participation diminishes their motivation to engage in NPRL activities.

*P5: “The main problem is that interdisciplinary consultations are with a limited number of therapists and GPs as time-investment and finance for these consultations are not anticipated*.*”* HCPs seek financial support from insurers for participating in these consultations. Changes in care organization, as proposed in NPRL, necessitate adjustments in reimbursement to facilitate collaboration.

*P30: I think in future, there will be finance to reimburse a practice nurses mental health for managing patients, for example in the eHealth application. If I need to do that myself, it will be difficult*.

##### Support structures: tools

Before participating in NPRL1.0, HCPs often based patient referrals solely on history and physical examinations, which made them feel unprepared for selecting, referring, and treating patients, leading to uncertainty.

Potential contributions of other HCPs in the treatment process were unclear, probably resulting in incorrect referrals and decreased quality of care. The eCoach-Pain introduced in NPRL1.0 aimed to improve collaboration but was found ineffective during NPRL2.0 focus groups due to time constraints and limited patient inclusion (Background information on the eCoach-Pain: (28)). A medical chat application used by therapists and mental health practice nurses, facilitated communication with GPs but was not integrated into the existing referral system resulting in calls for its broader adoption and standardization.

### Interprofessional Collaborative Practice

#### Collective ownership of goals: treatment

During NPRL1.0, HCPs appreciated being involved in the design and implementation process, which resulted in a shared vision on referral and treatment strategies and improved collaboration. Initially, uncertainties about NPRL’s development process arose, leading to ambiguity. As NPRL2.0 evolved, clarity increased, enhancing confidence in participation. The program facilitated pain education, leading to a shift in attitudes. HCPs preferred collective ownership and outcomes of biopsychosocial treatment over individual professional goals.

#### Collective ownership goals: participating in NPRL

In many local networks, GPs initially expressed interest in participating in NPRL and invited local therapists to join. However, most GPs did not actively participate in NPRL1.0 and NPRL2.0, resulting in disappointment among therapists. Therapists experienced challenges in collaboration with busy GPs and often turned to mental health practice nurses for cooperation instead. This posed issues as therapists required interdisciplinary consultations with GPs.

*P20: “The GP I collaborate with most recognized issues in usual CMP care but was too busy to participate in NPRL1*.*0. Too many hours*.*”*

HCPs indicated that healthcare centers with all disciplines involved were more successful than separate practices where engaging GPs was challenging.

Most HCPs reported no change in collaboration patterns due to NPRL2.0, especially in healthcare centers as the existing teams and collaboration structures were already effective for other diseases. They only used the existing teams and structures for the new CMP patients. Some therapists reported feeling unsupported by their colleagues, which complicated the situation. Seven HCPs deemed NPRL feasible in daily practice, but most felt more experience was needed for long-term improvement in collaboration.

Collaboration was generally perceived as good when HCPs shared a biopsychosocial treatment vision and goals, leading to uniform treatment plans. However, extensive time and energy were spent on interdisciplinary meetings and pain education implementation, while patient inclusion numbers remained lower than expected.

#### Knowledge exchange

At the start of NPRL1.0, HCPs aimed to conduct effective yet infrequent interdisciplinary consultations to keep motivation high, considering them essential for team success. Initially, collaboration was more intensive than in NPRL2.0. Due to difficulties in scheduling structural interdisciplinary meetings, the HCPs organized fewer ad-hoc meetings later on. Lack of time of therapists was not cited as a reason for limited meetings, but the availability of the GPs and the lack of clear guidance were. To overcome this in the future, HCPs plan to integrate NPRL into existing meetings for other diseases with multiple HCPs present.

Integrated network events were found to boost collaboration and patient referrals, with secondary and tertiary care HCPs referring more patients to primary care due to better knowledge of primary care treatment content. Primary care HCPs eagerly anticipate in-person meetings with all participants to share experiences and learn about secondary and tertiary care treatments. They also aim to educate non-participating HCPs about NPRL2.0 to improve collaboration and enhance referrals.

Some HCPs desire more support to enhance their CMP knowledge; they indicated that CMP and biopsychosocial model education would increase job satisfaction.

#### Understanding of roles

At the start of NPRL2.0, HCPs emphasized the importance of respecting each other’s strengths.

*P12: “I agree that you must have respect for each other’s strengths. You have to know them and use them. The healthcare practices in secondary and tertiary care have their strengths and expertise, which we do not have in primary care. In primary care, we have close patient contact, which is not the case in secondary and tertiary care*.*”*

In the future, HCPs aim to involve additional disciplines such as practice nurses in mental health or psychologists as case managers to provide comprehensive long-term care for patients-with CMP. They do not seek to increase the number of HCPs per local collaboration but rather focus on expanding the number of local collaborations for better regional coverage.

#### Work behaviors & attitudes

##### Work satisfaction

HCPs noted that the inability to treat patients with CMP sometimes resulted in stress, which reduced work satisfaction. They participated in NPRL to adopt a proactive approach in identifying patients with subacute pain or CMP, aiming to enhance their skills in early recognition of these patients and refer them to specialized colleagues within NPRL as needed.

P4: *“Sometimes, you have to show your vulnerabilities. If you cannot help a patient, but someone else has specific expertise, then I can refer the patient to that colleague*.*”*

Participation in NPRL1.0 and NPRL2.0 led to increased job satisfaction as HCPs experienced successful collaboration with colleagues from various disciplines. They found joy in collaborative treatment, gained inspiration, and felt stress reduction through discussions. Improved team effectiveness and patient care quality were perceived benefits.

However, administrative tasks in current care settings, outside NPRL, decreased joy in work. Some HCPs rearranged their practices to allocate more time for administrative tasks, even though this led to decreased patient care and income.

##### Work stress

Before the start of NPRL1.0, HCPs expected their stress levels to be stable but recognized the initial need for a significant investment of time. *P14: “In the beginning, it will take a larger time-investment. You have to be realistic. However, I do not know what the influence will be on my stress level [agreement with other participants]*.*”*

HCPs highlighted the challenge of short consultation times, leading to high time pressure. They hoped to allocate sufficient time for NPRL1.0.

By the end of NPRL2.0, seventeen HCPs highlighted the need to refine the content. Some participants found the assessment tool stressful and time-consuming which increased their stress level, further exacerbated by involvement in other demanding healthcare projects. This led to diminished engagement in NPRL. Ongoing participation still contributes to stress due to suboptimal implementation of tools and inadequate integration into clinical routines.

##### Perceived team effectiveness

Nine HCPs reported struggles with the limitations of biomedical perspectives held by non-participating HCPs and patients. Additionally, varying levels of knowledge about CMP among participating HCP disciplines decreased job satisfaction and hindered collaboration within and outside NPRL1.0 and NPRL2.0.

Therapists noted that while the assessment tool occasionally recommended referrals to secondary or tertiary care, their colleagues within their practice not participating in NPRL failed to see the value of this biopsychosocial approach and were concerned about potential income loss due to fewer consultations. Participating therapists felt their colleagues took them less seriously as a result.

#### Patient outcomes

##### Biopsychosocial outcomes

Patients with CMP frequently expected a biomedical approach, which posed challenges for HCPs attempting to implement a biopsychosocial approach. Even within NPRL, managing patients with CMP remained a complex task for HCPs.

##### Satisfaction

Before the start of NPRL1.0, HCPs viewed the initiative as trustworthy, with clear treatment plans for both patients and professionals that met their expectations. They viewed NPRL’s interdisciplinary approach as enhancing the value of usual care, giving patients the impression that they were receiving additional attention.

Within NPRL, five HCPs noted benefits for the patients which increased satisfaction. They indicated that patients felt that their complaints were taken seriously and accepted by their HCPs.

In future care, HCPs aim to enhance patient information delivery about NPRL’s goals to better prepare them for participation and a biopsychosocial approach.

#### Organizational outcomes

##### Length of treatment

In NPRL1.0, GPs noticed a shift in referral patterns. When they informed patients about the therapist’s treatment content, patients were better prepared and more willing to participate. Therapists found this added value and it energized their treatment delivery.

Despite financial challenges in organizing multidisciplinary meetings and insurance packages, HCPs anticipated future financial benefits by delivering timely and appropriate care. They expected time savings in the treatment process due to better-suited referrals. However, they emphasized the need for shorter waiting times in secondary or tertiary care to avoid hindering patients’ treatment plans.

##### Interprofessional Collaboration Competencies Attainment Survey

Of HCPs working in NPRL2.0 (n=37), 19 filled in the ICCAS (response rate of 51%); 15 participated in focus groups while four did not attend focus groups.

Before participating in NPRL2.0, HCPs perceived their ability to engage in ICP to be good (score of 3) or very good (score of 4) (Table 3). After participating in NPRL2.0 they perceived their ability to engage in ICP as very good (score of 4) on all constructs. Mean after-NPRL2.0 scores were higher than their before-NPRL2.0 for all constructs. There was a significant difference (p < 0.05) and a greater magnitude of change in the construct communication and team functioning. The effect size of ‘conflict management/resolution’ was negligible (<0.20), of ‘collaboration’ and ‘roles and responsibilities’ small (0.20<0.50), and ‘communication’ medium (0.50<0.80). Team Functioning had a large (>0.80) effect size as measured by Cohen’s d.

**Table 3.**
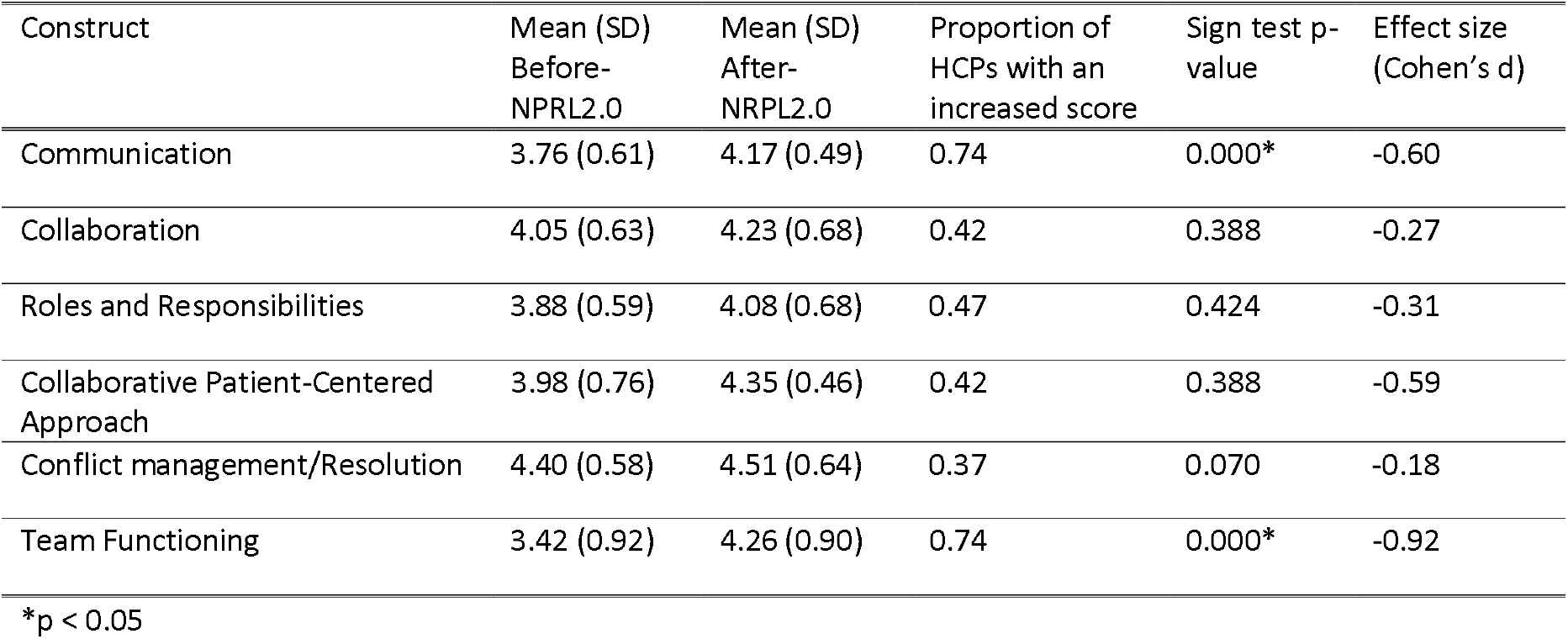
Comparison of before-NPRL2.0 and after-NPRL2.0 self-ratings for the ICCAS (n=19).

| Construct | Mean (SD)<br>Before-NPRL2.0 | Mean (SD)<br>After-NPRL2.0 | Proportion of<br>HCPs with an<br>increased score | Sign test p-<br>value | Effect size<br>(Cohen's d) |
| --- | --- | --- | --- | --- | --- |
| Communication | 3.76 (0.61) | 4.17 (0.49) | 0.74 | 0.000* | -0.60 |
| Collaboration | 4.05 (0.63) | 4.23 (0.68) | 0.42 | 0.388 | -0.27 |
| Roles and Responsibilities | 3.88 (0.59) | 4.08 (0.68) | 0.47 | 0.424 | -0.31 |
| Collaborative Patient-Centered<br>Approach | 3.98 (0.76) | 4.35 (0.46) | 0.42 | 0.388 | -0.59 |
| Conflict management/Resolution | 4.40 (0.58) | 4.51 (0.64) | 0.37 | 0.070 | -0.18 |
| Team Functioning | 3.42 (0.92) | 4.26 (0.90) | 0.74 | 0.000* | -0.92 |

\*p < 0.05

## Discussion

This study aims to describe the perceived ICP and work satisfaction of HCPs participating in a transmural network with a biopsychosocial approach. Overall, HCPs mentioned positive experiences but no main changes in ICP and work satisfaction were found. In NPRL, an integrated biopsychosocial rehabilitation for patients with CMP was introduced. However, more time seems to be needed to experience advantages in ICP and work satisfaction. NPRL2.0 is not embedded in such a way that it is used regularly. Further optimalization is needed. However, after participating in NPRL 2.0 a significant improvement and a greater magnitude of change in communication and team functioning was found compared to before the start of NPRL2.0.

### Personal

This study found that personal factors, such as motivation to participate, positively influenced ICP in NPRL 1.0 and 2.0, while situational factors related to the financial organization of healthcare had a negative impact on ICP. Before NPRL1.0 participation, HCPs often cited the need for improved communication and trust among peers. However, over time, HCPs reported more positive attitudes and increased engagement in ICP, consistent with previous research on interprofessional learning [39,40]. Furthermore, when encountering issues with the current organization of care for CMP, HCPs showed heightened willingness to participate.

### Situational

The financial structure of Dutch healthcare poses barriers to ICP and diminishes HCPs’ motivation for participation. Previous research demonstrated reduced referrals to secondary and tertiary care when GPs had more time per patient and were compensated per patient rather than per consultation [41]. Participation of GPs in multidisciplinary consultations fostered a more comprehensive biopsychosocial approach. Financing transmural care and interdisciplinary consultations in primary care could enhance ICP in the Netherlands and boost HCPs’ motivation for interdisciplinary collaboration. Szafran et al. (2018) identified appropriate payment mechanisms as facilitators for ICP [42]. In our study, other situational factors, such as the tools used in NPRL2.0, did not significantly improve ICP and team effectiveness, although they may have had benefits for individual HCPs in terms of treatment and confidence.

### Interprofessional Collaboration Practice

HCPs felt a strong sense of collective ownership (aligning values) regarding the goals of biopsychosocial treatment, recognizing its pivotal role in fostering successful ICP, as also highlighted by Wei et al [2022] [23]. During discussions on NPRL’s driving force for sustainable collaboration, it became evident that effective coordination by a project management group from a rehabilitation center specialized in pain rehabilitation treatment, alongside leadership from GPs in local networks, was crucial. However, organizing interdisciplinary meetings with GPs posed a significant challenge due to their busy schedules, resulting in ad-hoc and infrequent meetings. This issue, previously reported, underscores the struggle to secure active GP participation, essential for the success of ICP [43–45]. GPs involvement is crucial as they are identified as particularly effective champions, but they are also the most difficult group to reach [46]. To address this, suggestions were made to integrate meetings into existing structures and focus discussions on the biopsychosocial approach tailored to patients with CMP. Notably, such an approach is not limited to CMP alone but could be extended to benefit patients with other chronic conditions or potentially become chronic conditions, potentially enhancing knowledge exchange among HCPs. Szafran et al. (2018) highlighted various barriers such as limited space, staff turnover, and power dynamics within primary care teams, while also identifying communication, trust, clearly defined roles, co-location, task shifting, and appropriate payment mechanisms as facilitators for effective teamwork [42].

In the primary healthcare centers, the way of collaboration did not change as it were already existing teams. However, communication and team functioning improved, with moderate and large effect sizes, after participating in NPRL. It also had been previously shown that shared work spaces (co-location) can lead to effective communication when agreements are made about how and when to collaborate [23]. HCPs not working in healthcare centers were already glad that local networks were created and that team functioning improved, even though there was not an optimal organization of interdisciplinary meetings and time-investment of GPs.

### Work behaviors & attitudes

Regarding the Integrated ICP and Quadruple Aim framework (Figure 2), ICP results in work behaviors & attitudes, patient outcomes, and organizational outcomes. Work satisfaction was improved as HCPs enjoyed the collaboration, got inspiration for treatment content, and noticed stress reduction through discussions with colleagues. Effective interdisciplinary primary care leads to fewer burnouts and intentions to leave the current position [47]. In our study, stress was increased by the extra administration and time-pressure of the consultations with patients. Working with non-participating HCPs decreased work satisfaction as they had often another vision regarding CMP. It has been shown that common team goals and collective ownership of goals lead to increased work satisfaction [48–50]. To improve ICP and work satisfaction other Quadruple Aim outcomes must stay at least equal [14]. Therefore, it is expected that better health outcomes and improved patient satisfaction will also improve HCPs’ satisfaction.

### Patient outcomes

Some HCPs indicated that patients felt being taken more seriously and accepted. However, patients’ biomedical thoughts were still difficult to overcome by HCPs. It has been shown that patients struggle to be recognized as ill, and a mismatch exists between patients’ experience of CMP and the HCPs’ view which can lead to ineffective treatment [51,52].

### Organizational outcomes

The organizational structure of NPRL, in which patients were referred back to a GP before a therapist consultation, made patients better prepared for their treatment. At the moment, no financial benefits were seen by the HCPs for participating in multidisciplinary meetings, although they anticipate such reimbursements in future care [53]. Additionally, HCPs mentioned that employing a case-manager in primary care could reduce work pressure and stimulate interdisciplinary care. A case-manager, such as a practice nurse, could be one strategy to improve access, efficiency, and quality of care [54]. However, attention must be paid to interprofessional education for ICP between GPs and practice nurses [55].

### Strengths and limitations

This study differs from other studies mainly because of the mixed-methods with several data collection moments and the integrated interdisciplinary focus. Due to these collection moments over time, the same HCPs attended focus groups and interviews at different time points to see changes regarding ICP and work satisfaction. In this mixed-methods study qualitative-alongside quantitative data were gathered, which made it possible to study ICP and work satisfaction from different points of view. Moreover, a strength was that one individual researcher was available as an interviewer or observer during all measurements. All data were analyzed by two researchers, which increased internal consistency.

However, not all HCPs performed all measurements or participated during the whole study in NPRL. It was sometimes difficult to compare HCPs in different implementation stages at different time-points. Additionally, participation was voluntary for HCPs and may have led to selection bias (e.g. HCPs experiencing higher work-related stress because of time-pressure might not have participated, or, organizations with lower response rates might have had higher levels of work-related stress). Another limitation is that the ICCAS questionnaire used in this project has not yet been officially translated and validated into Dutch. Further research and a cross-cultural validation must be performed to confirm the translation and test the validity. Moreover, as the ICCAS asked HCPs retrospectively their perceived ability to engage in ICP, the long study period made it hard to recall their pre-NPRL experience, potentially affecting their scores. Consequently, this could have influenced their scoring on the ICCAS. The analysis of qualitative data also presented challenges. Due to the relatedness of concepts in the Integrated ICP and Quadruple Aim framework, the decision where some conclusions could be placed during analysis was sometimes difficult. A structured framework to assess and guide ICP does not exist yet [17,23]. Finally, the generalizability of the results to other countries may be limited, because of differences in the organization and financing of healthcare systems.

### Recommendations and conclusions

Based on the results of this study in patients with CMP, it can be concluded that while HCPs had positive experiences with interdisciplinary collaboration in primary care, there were no considerable changes in ICP and work satisfaction. There is a commitment to interdisciplinary collaborations in primary care to guide patients with CMP. It is recommended to continue this integrated rehabilitation treatment for patients with CMP in a new version of NPRL: NPRL3.0. In the next steps, attention should be given to the barriers found in this study to improve ICP and increase work satisfaction. Subjects of improvement could be: promoting a more biopsychosocial view of society regarding CMP, the introduction of a case-manager in primary care to unburden general practitioners, and a different way of reimbursement of HCPs by health insurers. The expectation is that the benefits in terms of ICP and work satisfaction will become increasingly visible and more and more experienced as a result.

## Data Availability

All data produced in the present study are available upon reasonable request to the authors.

## Acknowledgments

We would like to thank Gijs Brouwer (G.B.), for leading the focus groups of NPRL1.0 and Petra de Koning (P.K.) for observing of a focus group in NPRL2.0. Furthermore, we would like to thank Eline Willems (E.W.) and Anouk Klinkenberg (A.K.) for their roles in the focus groups of NPRL2.0 as part of their master thesis.

## Data availability statement

The data that support the findings of this study are available on request from the corresponding author, C.L. The data are not publicly available due to restrictions, their containing information that could compromise the privacy of research participants.

## Author Contributions statement

C.L. was involved in conception and design, analysis and interpretation of the data and drafting of the paper, revising it critically for intellectual content. M.K, J.V. and I.H. were involved in supervision, design, interpretation of the data, and writing—review & editing. M.d.M. was involved in formal analysis, investigation, validation, and revising it critically for intellectual content. All the authors meet the criteria for authorship as per the ICMJE criteria and they have read and agreed to the published version of the manuscript.

