## Supplementary material for "Healthcare professionals’ collaboration and satisfaction within an innovative primary care network for patients with chronic musculoskeletal pain: a mixed method study": S2

### S2 Topic list focus groups and interviews

#### Focus groups 1&2 – primary care

- General opinion about NPRL
    - Adequate level of knowledge and resources
    - Added value for daily practice
    - Useability of tools

- Start meeting
    - Content
    - Alignment with daily practice
    - Way healthcare professionals are involved in NPRL

- Education meetings
    - Usefull for daily practice
    - Knowledge about CMP

- Practice meetings
    - Added value
    - Implementation of disucced topics in daily practice

- Assessment tool 1
    - Usability
    - Practical use
    - Complexity of patients
    - Integration in eHealth

- eHealth
    - Usability
    - Practical use
    - Comparison with other eHealth applications
    - Added value

- Treatment protocol
    - Usability
    - Fixed protocol in the future
    - Adaptations in protocol during education meetings

- Collaboration
    - Interdisciplinary collaboration
    - Subdividing tasks
    - Difference with usual care

- Expectations for the future
    - Implementation of NPRL in daily care
    - Implementation in your practice/organisation
    - Barriers

**Focus group 3 – primary care**

- Additional education recognition patient with CMP
    - Usability
    - Understandable
    - Added value for use in daily care
    - Need for extra education/information

- Collaboration
    - Workshops from secondary and tertiary care
        - Understandable
        - Referral to secondary and tertiary care
        - Need for extra information
    - Interdisciplinary collaboration in local network
        - Use of practice nurse mental health

- Treatment protocol
    - Summary of the protocol
    - Need for extra education/information

- eHealth
    - Extra education
    - Usability daily practice
    - Need for extra education/information
    - Preventive care

- Assessment tool
    - Usability extra rules and information
    - Integration in daily care
    - Need for extra education/information

- Network meeting with all healthcare professionals
    - General opinion
    - Points for improvement
    - Subject next edition
    - Summary of the network meeting

- Participation of patients
    - Eligible patients who did not start treatment
    - ‘Automatic process’ of inclusion and treatment

- Expectations for the future
    - What do you need?
    - Barriers
    - Adjustments

**Interview 1 - practice nurse mental health – primary care**

- Network meeting with all healthcare professionals
  - General opinion
  - Points for improvement

- Collaboration
  - Facilitators
  - Local network
  - Comparison with usual care

- Treatment protocol
  - Specific for practice nurse mental health
  - Content

- eHealth
  - Experiences
  - Feedback system
  - Collaboration

- Participation of patients
  - Financial situation
  - Satisfaction

- Transferability of NPRL
  - Healthcare disciplines
  - Local network

**Focus group 4 – primary, secondary and tertiary care**

- Experiences in phase 2
    - Barriers
    - Facilitators

- Network meeting with all healthcare professionals
    - General opinion
    - Points for improvement
    - Subject next edition
    - Summary

- Collaboration
    - Interdisciplinary collaboration
    - Local network
    - Local networks vs secondary/tertiary care
    - eHealth and collaboration

- Network vs. usual care
    - Activities
    - Way of working

- Participation of patients
    - 'Automatic process' of inclusion and treatment
    - Barriers
    - eHealth and participation
    - Biopsychosocial model

- Assessment tool 2
    - New version
    - Usability
    - Added value

- Expectations for the future
    - Treatment of all available patients in NPRL
    - Barriers
    - Adjustments
    - Continuity NPRL inside each organization/ practice
    - Education meetings

**Focus group 5, 6 and 7 – primary care**

Introduction: 10 min
- Introduce yourself
- ☐ Function
- ☐ Background
- ☐ Practice / discipline
- ☐ How did you get involved?

Tools (ECoach Pain Rehabilitation, assessment tool, treatment protocols etc.): 20 min
- Which tools did you get within NPRL?
- Which tools did you use? How did you use this? With which purpose?
- Do you have the idea that these tools work? And why?
- How often do you use these tools?
- What is your opinion about the use of these tools and the time investment?

Currently, there is a new version of the assessment tool developed, translated and validated in the Netherlands.
- What is your opinion about this new version?
- Do you have the idea you can use this new version in daily practice?
- Could this version have an added value for your workload?

Also the treatment protocols are updated for treatments with blended care on the long term.
- What is your opinion about this new version?
- Do you have the idea you can use this new version in daily practice?
- Could this version have an added value for your workload?
- Do you think telemedicine can be of added value for your work?
- Do you want to use the telemedicine options?
- How do you want to get informed about the new version of the treatment protocol? Do you want extra training, information leaflets or …?

Education: 20 min
- What is your opinion about the training you received for NPRL? (informative, use in practice etc.)
- What is your experience with giving pain education after following the training?
- How do you incorporate the information of the training in your work?
- Do you experience a difference in giving pain education?
- Did the training help you in treating patients?
- Did the training had an influence at your work satisfaction?
- Do you have the feeling you need more training?

Experiences with working in NPRL: 10 min
- How many patients with subacute and chronic complaints did you see last week?
- Do you have the idea that you treat enough patients to optimal integrate NPRL in daily practice?
- Did the work pressure change due to NPRL? And how?
- Do you have the idea that you can adequately help the patients?
- Is there a respectfull relation between the patient and HCP?
- Do you experience a difference in contact with patients since participation in NPRL?

ICP: 10 min
- How do you exprience the collaboration between HCPs in NPRL?
- How do you exprience the collaboration between HCPs of different disciplines in NPRL?
- Was the collaboration respectfull?
- Where there barriers in ICP? How did these barriers look like? Did you found a solution for these barrriers?
- How is the collaboration between the physiotherapists and general practitioners?
- How often is their contact with other HCPs?
- Did you experience any change because of the collaboration in NPRL?

Meaning & Joy: 10 min
- To which extent has NPRL influence at your work pleasure?
- To which extent has NPRL influence at your meaning in work?
- To which extent has NPRL influence at your joy in work?
- What could or must be changed to increase joy and meaning in work for NPRL?

Stress: 5 min
- Do you have work stress when treating patients with CMP? How often?
- Does NPRL has influence at your level of work stress? How?
- What could or must be changed to work stress because of NPRL?

Improvements: 10 min
- What do you expect of NPRL in the coming year?
- What do you want to learn in the coming year?
- Which improvements would you suggest for NPRL?

**Focus groups 8, 9, 10 and 11 – primary care**

Introduction – 5MIN
- Introduce yourself
- Function
- Background
- How did you, so far, experience working NPRL?
- Is your vision about the treatment of patients with CMP changed due to participation in NPRL? How was it before participation? What do you think of this change?

Content of NPRL – 15 MIN
Education and training
- How did you experience the education and training received by NPRL?
- What is your experience with giving pain education after following the training?
- How do you incorporate the information of the training in your work?
- Do you experience a difference in giving pain education?
- Did the training help you in treating patients?
- Did the training had an influence at your work satisfaction?
- Do you have the feeling you need more training?

Assessment tool
- What is your opinion about this new version?
- Do you have the idea you can use the assessment tool daily practice?
- Could this version have an added value for your workload?
- Does the assessment tool help in assessing the complexity of patients' complaints?

Treatment protocol
- What is your opinion about the treatment protocol?
- Do you have the idea you can use this new version in daily practice?
- Could this version have an added value for your workload?

Tools (ECoach Pain Rehabilitation, assessment tool, treatment protocols etc.): 20 min
- Which tools did you get within NPRL?
- Which tools did you use? How did you use this? With which purpose?
- Do you have the idea that these tools work? And why?
- How often do you use these tools?
- What is your opinion about the use of these tools and the time investment?

Experiences with working in NPRL: 10 min
- How many patients with subacute and chronic complaints did you see last week?
- Do you have the idea that you treat enough patients to optimal integrate NPRL in daily practice?
- Did the work pressure change due to NPRL? And how?
- Do you have the idea that you can adequately help the patients?
- Is there a respectfull relation between the patient and HCP?
- Do you experience a difference in contact with patients since participation in NPRL?

ICP: 30 min
- How do you exprience the collaboration between HCPs in NPRL? Did it change since the start of NPRL?
- How do you exprience the collaboration between HCPs of different disciplines in NPRL? Did it change since the start of NPRL?
- Was the collaboration respectfull? Did it change since the start of NPRL?
- Where there barriers in ICP? How did these barriers look like? Did you found a solution for these barrriers?
- How is the collaboration between the physiotherapists and general practitioners? Did it change since the start of NPRL?
- How often is their contact with other HCPs?

Communication:
- How did you experience the communication with HCPs of NPRL? Change?
- How did you experience the communication with HCPs outside NPRL? Change?

- Do you want to improve the communication? How? Why? Respectfull?

Tasks
- Do all HCPs have a clear role in NPRL? Did this change since the start?
- Do you have the idea that all HCPs have the same goal?
- Do you have the idea that you are respected by the other HCPs?
- Change since the start?

Expectations
- Does the vision of NPRL about ICP has influence the way of collaboration between the HCPs?
- If not, what do you need to change this?
- Had ICP an influence at the way you refer or treat patients with CMP? Example?
- Is ICP of added value for you?

SITUATIONAL factors – 10MIN
- Is there a clear leader, project group, or someone who facilitates NPRL?
- Do you have the idea that you have enough time in your work schedule for NPRL? And contact with other HCPs?
- Does your work enviroment facilitate participation and collaboration in NPRL? Why?

Quality of care - 15MIN
- What is, for you, the added value of delivering of care following the principles of NPRL? Examples?
- What do you think about the quality of care for patients with CMP in NPRL? Barriers? Facilitators? Added value – care as usual?
- What is, in your opinion, the added value for patients? Satisfied patients?
- Do you see more patients with CMP since pariticaption in NPRL? Treatment more adequate? More or less consultations necessary?

WORK BEHAVIOURS & ATTITUDES – 15 MIN
Meaning & Joy- To which extent has NPRL influence at your work pleasure?
- To which extent has NPRL influence at your meaning in work?
- To which extent has NPRL influence at your joy in work?
- What could or must be changed to increase joy and meaning in work for NPRL?

Stress:
- Do you have work stress when treating patients with CMP? How often?
- Does NPRL has influence at your level of work stress? How?
- What could or must be changed to work stress because of NPRL?

SUMMARY – 5MIN
- What is your point of view about: NPRL is of added value compared to care as usual for patients with CMP.
- What is your point of view about: NPRL is, for me as HCP, feasible in daily practice.

- What do you expect of NPRL in the coming year?
- What do you want to learn in the coming year?
- Which improvements would you suggest for NPRL?
