## Supplementary material for "Healthcare professionals’ collaboration and satisfaction within an innovative primary care network for patients with chronic musculoskeletal pain: a mixed method study": S3

**Supplementary file 3**

These concepts used in the framework of this study were based on the Conceptual Framework for Interprofessional Collaborative Practice, Job-Demand-Control-Support Model, and Quadruple Aim [16-18].

| Concept | Definition |
|---|---|
| ***Personal Factors*** | ***Controlled internally by an individual*** |
| Beliefs in Interprofessional Collaboration | Strengths and weaknesses identified by HCPs in interprofessional collaboration in NPRL as well as their beliefs for the future. |
| Flexibility | Deliberate role-blurring including reaching productive compromises |
| Trust | The confidence and reliance that HCPs have with their own and others' competencies and NPRL |
| Cooperation | The manner in which HCPs work together for a common goal and are open for collaboration |
| Communication skills | The ease and effectiveness with which professionals communicate with each other |
| Control | Employees' freedom to use specific job skills at work, and employees' autonomy in task-related activities. This concerns the freedom an employee has to control and organise his own work. This latitude refers to the control that employees have about their duties and how they want to perform these tasks. It consists of both competence and decision-making authority. |
| ***Situational Factors*** | ***Factors that HCPs are exposed to within the workplace that either support or interprofessional collaboration*** |
| Leadership | Both central and local leadership to promote collaboration, eliminate barriers and promote an effective team culture, where local is their own practice/workplace and central the project team coordinating the entire NPRL |
| Support Structures | Having the physical space, time, policies and procedures, and formal mechanisms to support interprofessional collaborative practice. Includes having adequate time for sharing knowledge and patient-related information and integrating daily collaborative behaviours into day-to-day functioning. Also consists of emotional support, helpful advice, or hands-on assistance from superiors, peers and interprofessional practitioners |
| Empowerment | Empowering environment including having access to information, support, resources, and the opportunity for growth and mobility |
| Demands | Psychological stressors involved in accomplishing the workload. These are the requirements that are set at work, including work rate, availability, time pressure, effort and difficulty. Such requirements represent the psychological stressors in the work environment. |

| | |
|---|---|
| ***Interprofessional Collaborative Practice*** | ***Multiple HCPs from different backgrounds provide interdisciplinary treatment by working with patients to deliver the highest quality of care across settings*** |
| Collective Ownership of Goals | Shared responsibility in the entire process of reaching goals, including joint design, definition, development, and achievement of goals and includes commitment to patient-centered care whereby HCPs from different disciplines and patients are all active in the process of goal attainment |
| Knowledge Exchange | Perception of the extent to which knowledge is shared between HCPs in local networks NPRL |
| Understanding of Roles | HCPs' knowledge and understanding of their role and the roles of other HCPs within NPRL |
| Interdependence | The occurrence of, and reliance on, interactions among HCPs whereby each is dependent on the other to accomplish his or her goals and tasks |
| ***Work Behaviours & Attitudes*** | ***Work behaviour in general and attitudes towards ICP as organised in NPRL. Both personal and team in nature*** |
| Work Satisfaction (joy & meaning) | Overall satisfaction with work in general and in NPRL |
| Work stress | When demands are high, control and support is low, HCPs are more prone to develop work stress. |
| Motivation | A reason for participating in NPRL. |
| Intent to Stay | Intent to stay in one's current job and NPRL |
| Perceived Team Effectiveness | The perceived effectiveness of the team of HCPs in NPRL concerning the ability to meet patient (and family) care needs and outcomes |
| Conflict | The degree to which HCPs disagree over the sharing of responsibilities and (group)decisions as well as general issues affecting NPRL |
| Support | Overall levels of helpful social interaction available on the job from both co-workers and supervisors. |
| ***Patient Outcomes*** | ***Outcomes of NPRL which are important for participating patients.*** |
| Biopsychosocial outcomes | The biopsychosocial model is a general model positing that biological, psychological (which includes thoughts, emotions, and behaviors), and social (e.g., socioeconomical, socioenvironmental, and cultural) factors, all play a significant role in health and disease. |
| Satisfaction | The extent to which a patient is content with the health care which they received from their health care provider. |
| Empowerment | A process that helps patients gain control over their own lives and increases their capacity to act on issues that they themselves define as important. |
| ***Organizational Outcomes*** | ***Outcomes of NPRL on the level of the organization of care.*** |
| Length of treatment | Duration and frequency of consultation and total treatments. |
| Quality of patient care | The degree to which health services for individuals and populations increase the likelihood of desired health outcomes. |
| Patient safety | The prevention of errors and adverse effects to patients associated with health care |
