## Supplementary material for "Healthcare professionals’ collaboration and satisfaction within an innovative primary care network for patients with chronic musculoskeletal pain: a mixed method study": S1

### Supplementary file 1

**Table 1: Overview of participating healthcare professionals in the various parts of the study.**

| | | | | | December 2017 | | May 2018 | | October 2018 | April 2019 | | | April 2020 | | | | April 2020 |
|---|---|---|---|---|---|---|---|---|---|---|---|---|---|---|---|---|---|
| | | | | | FG 1 | FG 2 | FG 3 | INT 1 | FG 4 | FG 5 | FG 6 | FG 7 | FG 8 | FG 9 | FG 10 | FG 11 | Q 1 |
| Duration (hours) | | | | | 1:45 | 1:26 | 1:35 | 0:27 | 1:34 | 0:55 | 0:58 | 0:32 | 1:13 | 2:02 | 1:10 | 1:15 | |
| | Discipline | Gender | Practice number | Years experience | | | | | | | | | | | | | |
| P1 | PT | F | 1 | 2.5 | x | | x | | | | | | | | | x | x |
| P2 | PT | M | 1 | 33 | | x | | | | | | | | | | | |
| P3 | PT | M | 2 | 34 | x | | x | | | x | | | | | | | |
| P4 | PT | M | 2 | 7 | | | | | | x | | | | | | | |
| P5 | PT | M | 12 | 38 | x | | | | | | | | | | | | |
| P6 | PT | M | 3 | 0.5 | x | | | | x | x | | | | | | | |
| P7 | PT | F | 3 | 7 | | x | x | | | | | | | | | | |
| P8 | PT | F | 4 | 0.5 | | | | | | | | | | x | | | x |
| P9 | PT | M | 4 | 22 | | | | | | | x | | | | | | x |
| P10 | PT | M | 5 | 20 | | | | | | | | x | | x | | | x |
| P11 | PT | F | 6 /7 | 20 | | | | | | | | x | x | | | | x |
| P12 | PT | M | 6 /7 | 36 | | | | | | | | x | x | | | | x |
| P13 | PT | M | 8 | 30 | | x | | | | | | | | | | | |
| P14 | PT | F | 9 | 34 | | | | | | | | x | | | | | |
| P15 | PT | F | 10 | | | | | | | | | | x | | | | x |
| P16 | PT | F | 10 | 4 | | | | | | | | | x | | | | x |
| P17 | PT | M | 10 | | | | | | | | | | | | | | x |
| P18 | PT | F | 11 | 1 | | | | | | | | | | | x | | x |
| P19 | PT | M | 11 | 20 | | | | | | | | | | | x | | x |
| P20 | ET | F | 2 | 25 | | x | x | | | x | | | | | | | x |
| P21 | OT | F | 4 | 17 | | | | | | | x | | | | | | |
| P22 | OT | F | 6 | 0.75 | | | | | | | | x | x | | | | x |
| P23 | OT | F | 6 | 20 | | | | | | | | | x | | | | x |
| P24 | OT | F | 9 | 3 | | | | | | | | x | | | | | |
| P25 | PNMH | F | 1 | 5 | | | | | | | | | | x | | | x |
| P26 | PNMH | F | 2 | | | | | x | | | | | | | | | |
| P27 | PNMH | F | 4 | 5 | | | | | | | x | | | | | | |
| P28 | PNMH | F | 4 | 10 | | | | | | | x | | | | | | |
| P29 | PNMH | F | 5 | 4 | | | | | | | | | | | | | x |
| P30 | GP | M | 1 | 10 | x | | x | | | x | | | | x | | | x |
| P31 | GP | F | 1 | | | | | | | x | | | | | | | |
| P32 | GP | M | 2 | 31 | x | | | | | | | | | | | | |
| P33 | GP | M | 2 | | | | | | | | | | | | | | x |
| P34 | GP | M | 3 | 8 | | x | | | x | | | | | | | | |
| P35 | GP | M | 4 | 17 | | | | | | | X | | | | | | |
| P36 | GP | F | 5 | 9 | | | | | | | | | | x | | | x |
| P37 | GP | F | 6 | | | | | | | | | | | | | x | |

FG: focus group; INT: interview; Q: questionnaire; EXP: Years experience in current job at first moment of participation; PT: physiotherapist; ET: exercise therapist; PNMH; practice nurse mental health; GP: general practitioner; PNT: patient; F: female; M: male; -: unknown; n.a.: not applicable
